# Super-spreading events initiated the exponential growth phase of COVID-19 with ℛ_0_ higher than initially estimated

**DOI:** 10.1101/2020.04.26.20080788

**Authors:** Marek Kochańczyk, Frederic Grabowski, Tomasz Lipniacki

**Affiliations:** Department of Biosystems and Soft Matter, Institute of Fundamental Technological Research, Polish Academy of Sciences, 02-106 Warsaw, Poland; Faculty of Mathematics, Informatics and Mechanics, University of Warsaw, 02-097 Warsaw, Poland

## Abstract

The basic reproduction number ℛ_0_ of the coronavirus disease 2019 has been estimated to range between 2 and 4. Here we used a SEIR model that properly accounts for the distribution of the latent period and, based on empirical estimates of the doubling time in the near-exponential phases of epidemic progression in several locations, we estimated that ℛ_0_ lies in the range 4.7–11.4. We explained this discrepancy by performing stochastic simulations of model dynamics in a population with a small proportion of super-spreaders. The simulations revealed two-phase dynamics, in which an initial phase of relatively slow epidemic progression diverts to a faster phase upon appearance of infectious super-spreaders. Early estimates obtained for this initial phase may suggest lower ℛ_0_.

## Introduction

The basic reproduction number ℛ_0_ is a critical parameter characterising the dynamics of an outbreak of an infectious disease. By definition, ℛ_0_ quantifies the expected number of secondary cases generated by an infectious individual in an entirely susceptible population. ℛ_0_ may be influenced by natural conditions (such as seasonality) as well as socioeconomic factors (such as population density or ingrained societal norms and practices) [1]. Accurate estimation of ℛ_0_ is of crucial importance because it informs the extent of control measures that should be implemented to terminate the spread of an epidemic. Also, ℛ_0_ determines the immune proportion *f* of population that is required to achieve herd immunity, *f* = 1 – 1/ℛ_0_.

A preliminary estimate published by the World Health Organization (WHO) suggested that ℛ_0_ of coronavirus disease 2019 (COVID-19) lies in between 1.4 and 2.5 [2]. Later this estimate has been revised to 2–2.5 [3], which is broadly in agreement with numerous other studies that, based on official data from China, implied the range of 2–4 (see, *e*.*g*., Liu *et al*. [4] or Boldog *et al*. [5] for a summary). This range suggests an outbreak of a contagious disease that should be containable by imposition of moderate restrictions on social interactions. Unfortunately, moderate restrictions that were implemented in, *e*.*g*., Italy or Spain turned out to be insufficient to prevent a surge of daily new cases and, consequently, nationwide quarantines had to be introduced.

We estimated the range of ℛ_0_ of COVID-19 based on the doubling times observed in the exponential phases of the epidemic in China, Italy, Spain, France, United Kingdom, Germany, Switzerland, and New York State. For each of these locations, we used trajectories of both cumulative confirmed cases and deaths [6]. Since our stochastic simulations suggested that the epidemic may have two-phase dynamics — slow (and susceptible to extinction) before any super-spreading events occur and fast and steadily expanding after the occurrence of super-spreading events — to capture the second phase of the trajectories, we analysed them after a fixed threshold of cases or deaths has been exceeded, in two-week intervals. Both the stochastic simulations and ℛ_0_ estimates were obtained within a susceptible–exposed–infected–removed (SEIR) model that correctly reproduces the shape of the latent period distribution and yields a plausible mean generation time. We concluded that the range of ℛ_0_ is 4.7–11.4, which is considerably higher than most early estimates. We conjecture that these early estimates were obtained for the first phase of the epidemic in which super-spreading events were absent.

## Results

### The SEIR model

We used a SEIR model (see Methods for model equations and justification of parameter values) in which:

- we assumed that the latent period is the same as the incubation period and is Erlang-distributed with the shape parameter *m* = 6 and the mean of 5.28 days = 1/*σ* [7];
- we assumed that the infectious period is Erlang-distributed with the shape parameter *n* = 1 (exponentially distributed) or *n* = 2, and the mean of 2.9 days = 1/*γ* [8, 9];
- the infection rate coefficient *β* was determined from *σ, γ, m, n*, and doubling time 𝒯_d_, which in turn was estimated based on the epidemic data as described in the next subsection, ultimately allowing us to estimate ℛ_0_ = *β*/*γ* as ℛ_0_(𝒯_d_).

The use of the Erlang distributions directly translates to the inclusion of multiple consecutive substates in the SEIR model, meaning that we assumed *m* ‘exposed’ substates and *n* ‘infectious’ substates (Erlang distribution is a distribution of a sum of independent exponentially distributed variables of the same mean).

### Estimation of ℛ_0_ in the exponential growth phase

First, we estimated the doubling time 𝒯_d_ within two-week periods beginning on the day in which the number of confirmed (in the SEIR model naming convention, ‘removed’, see Methods) cases exceeded 100 or the number of deaths exceeded 10 in China, six European countries, and New York State (figure 1*a* and figure 1*b*). Values of 𝒯_d_ that we obtained lie in between 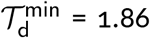 (based on cases in New York State) and 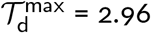 (based on deaths in Switzerland).

**Figure 1:**
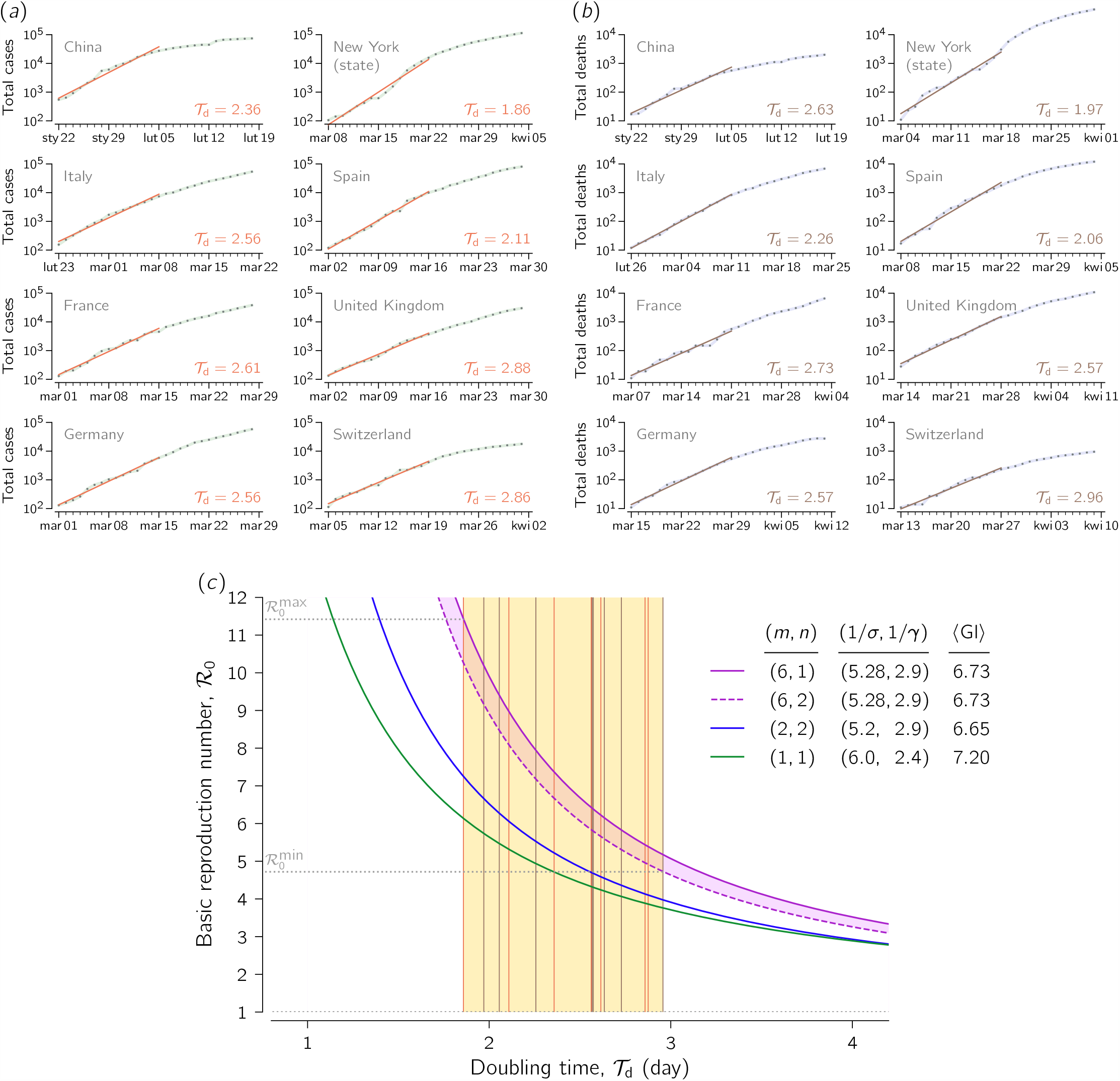
Estimation of the doubling time and the resulting basic reproduction number ℛ_0_. (*a, b*) Estimates of the doubling time 𝒯_d_ for China, six European countries, and New York State using two-week periods beginning (*a*) when the number of confirmed cases exceeds 100 or (*b*) when the number of deaths exceeded 10, according to data gathered and made available by Johns Hopkins University [6]. (*C*) The range of ℛ_0_ estimated using two variants of our SEIR model (violet solid and dashed curves) for the range of 𝒯_d_ estimated in panels *a* and *b*. Vertical lines in the yellow area are 𝒯_d_ estimates based on the cumulative number cases (orange, from panel *a*) or the cumulative number of deaths (brown, from panel *b*). Blue and green solid curves correspond to ℛ_0_(𝒯_d_) according to SEIR models structured and parametrised as in the study of Kucharski *et al*. [9] (*m* = 2, *n* = 2) and Wu *et al*. [14] (*m* = 1, *n* = 1).

Then, we estimated the range of ℛ_0_ as a function of the doubling time 𝒯_d_ using a formula that takes into account the mean latent and infectious period, 1/*σ* and 1/*γ*, respectively, as well as the shape parameters *m* and *n*, see equation (8) in Methods. The lower bound has been obtained using the model variant with *n* = 2 (two ‘infectious’ substates), whereas the upper bound results from the model with *n* = 1 (one ‘infectious’ substate), figure 1*c*. After plugging 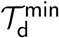 and 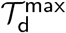 in, respectively, the variant of our model with the lower ℛ_0_(𝒯_d_) curve (*n* = 2) and the variant with the higher ℛ_0_(𝒯_d_) curve (*n* = 1), we arrived at the estimated ℛ_0_ range of 4.7–11.4. The cases-based doubling time for China, 2.34, is consistent with the value of 2.4 reported by Sanche *et al*. [10], who estimated that ℛ_0_ for China lies in the range 4.7 to 6.6, that overlaps with our estimated range for China: 5.6–7.3. The models having one or two ‘exposed’ substates, often used to estimate the value of ℛ_0_, substantially underestimated ℛ_0_, *cf*. figure 1*c* and the articles by Wearing *et al*. [11], Wallinga & Lipsitch [12], and Kochańczyk *et al*. [13].

There are two main reasons why our estimates of the basic reproduction number are higher compared to other published estimates:

1. Our SEIR model comprises 6 ‘exposed’ substates to account for the latent period distribution. As shown in figure 1*c*, broader latent period distributions, exponential or Erlang with *m* = 2, result in lower ℛ_0_ estimates (at the same remaining model parameters). We demonstrated sensitivity of ℛ_0_ with respect to the mean latent period, 1/*σ*, in electronic supplementary material (figure S1).
2. We estimated the doubling time, 𝒯_d_, from the growth of the number of cumulative cases and cumulative deaths in the two-week-long exponential phases of the epidemic in six locations, obtaining 𝒯_d_ ranging from 1.86 to 2.96. These values are much lower than the values reported in the early influential studies of Wu *et al*. [14, 15] and Li *et al*. [16]: 5.2 days, 6.4 days, and 7.4 days, correspondingly. In these studies the basic reproduction number has been estimated to lie in between 1.94 and 2.68. A summary in Table 1 shows that the lower ℛ_0_ estimates follow from much longer estimates of *T*_d_.

### Impact of super-spreading on 𝒯_d_ estimation

The discrepancy in 𝒯_d_ estimation may be potentially attributed to the fact that not all ‘removed’ individuals are registered. In the case when the ratio of registered to ‘removed’ individuals is increasing over time, the true increase of the ‘removed*’* cases may be overestimated. We do not rule out this possibility, although we consider it implausible as the expansion of testing capacity in considered countries has been slower than the progression of the outbreak. We rather attribute the discrepancy to the fact that in the early phase, in which the doubling time (growth rate) is estimated based on individual case reports, the consequences of potential super-spreading events (such as football matches, carnival fests, demonstrations, masses, or hospital-acquired infections) are negligible due to a low probability of such events when the number of infected individuals is low. In a given region or country, occurrence of first super-spreading events triggers transition to the faster-exponential growth, in which subsequent super-spreading events become statistically significant and may become decisive drivers of the epidemic spread [18]. Based on case reports in China, Sanche *et al*. [10] inferred that the initial epidemic period in Wuhan has been dominated by simple transmission chains. Phylogenetic analyses by Worobey *et al*. [19] revealed that first cases recorded in USA and Europe did not initiate sustained SARS-CoV-2 transmission networks. In turn, super-spreading events were very likely the main drivers of the epidemic spread in, *e*.*g*., Italy and Germany, where, in the early exponential phase, spatial heterogeneity of registered cases has been evident [20, 21]. In Italy, Spain, and France, this explosive phase was followed by a phase of slower growth, during which mass gatherings were forbidden, but quarantine (that finally brought the effective reproduction number below 1) has not been yet introduced.

**Table 1:**
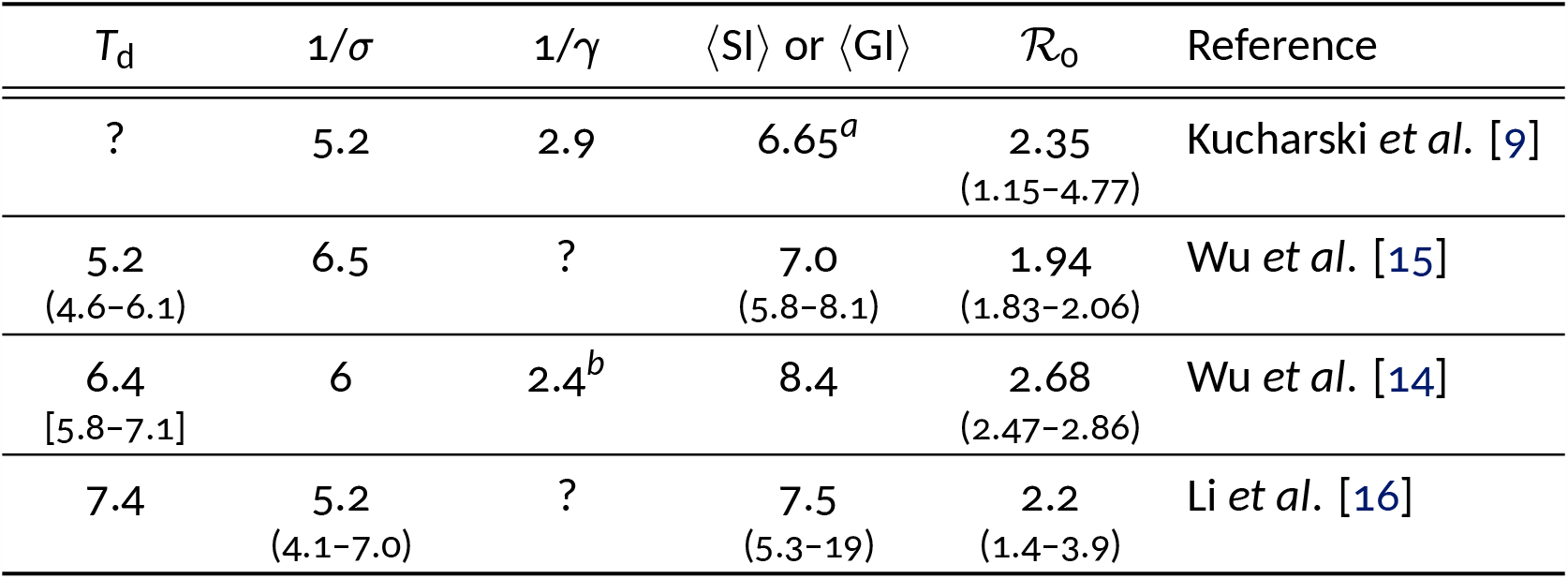
Relation between 𝒯_d_, model parameters (mean latent period or mean incubation period, 1/*σ*, mean period of infectiousness, 1/*γ*, and consequent mean generation interval, ⟨GI⟩), mean searial period, ⟨SI⟩, and ℛ_0_. The unit of all values, except for ℛ_0_, is day. Confidence intervals are given in oval brackets; a credible interval is given in square brackets. ^*a*^The ⟨GI⟩ value is not given in the article but calculated from the assumed values of 1/*σ* and 1/*γ* as 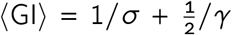 [17]. ^*b*^The value 1/*γ* was obtained by the authors as ⟨SI⟩ – 1/*σ*, which is inconsistent with the assumption that the infection occurs in a random time during the period of infectiousness.

| $\mathcal{T}_d$ | $1/\sigma$ | $1/\gamma$ | $\langle SI \rangle$ or $\langle GI \rangle$ | $\mathcal{R}_0$ | Reference |
| --- | --- | --- | --- | --- | --- |
| ? | 5.2 | 2.9 | 6.65<sup>a</sup> | 2.35<br>(1.15–4.77) | Kucharski *et al.* [9] |
| 5.2<br>(4.6–6.1) | 6.5 | ? | 7.0<br>(5.8–8.1) | 1.94<br>(1.83–2.06) | Wu *et al.* [15] |
| 6.4<br>[5.8–7.1] | 6 | 2.4<sup>b</sup> | 8.4 | 2.68<br>(2.47–2.86) | Wu *et al.* [14] |
| 7.4 | 5.2<br>(4.1–7.0) | ? | 7.5<br>(5.3–19) | 2.2<br>(1.4–3.9) | Li *et al.* [16] |

Motivated by these considerations, we analysed the impact of super-spreading on estimation of 𝒯_d_ based on stochastic simulations of SEIR model dynamics (see electronic supplementary material, listing S1). Simulations were performed in the perfectly mixed regime according to the Gillespie algorithm [22]. We assumed that a predefined fixed proportion of individuals (equal 33%, 10%, 3% or 1%) has higher infectiousness and as such is responsible for on average either half of infections (‘superspreaders’) or two-third of infections (‘hyper-spreaders’). To reproduce these fractions in systems with different assigned proportions of super- or hyper-spreaders, their infectiousness is assumed to be inversely proportional to their ratio in the simulated population. In figure 2 we show dynamics of the epidemic spread in the presence of 1% of hyper-spreaders to demonstrate that the phase of slower growth is transformed into the faster-exponential growth phase upon the occurrence of hyper-spreading events.

**Figure 2:**
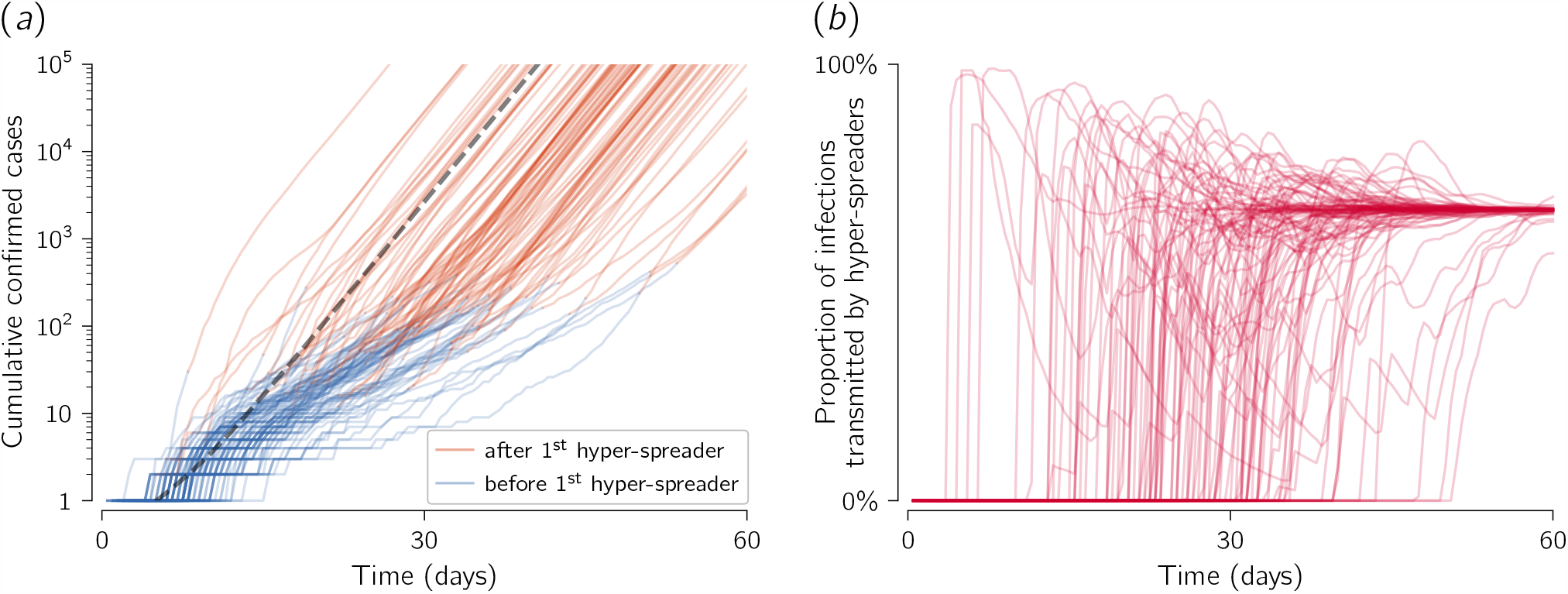
Stochastic epidemic spread in the presence of 1% of hyper-spreaders. (*a*) Trajectories of confirmed cases (cumulative *R* in terms of SEIR compartments) resulting from 100 independent stochastic simulations. When the first hyper-spreading event occurs, the colour of the line is changed from blue to brown. Dashed grey line shows a deterministic trajectory. (*b*) Proportion of infections transmitted by hyper-spreaders among all transmission events over time. Stochastic trajectories stabilise at 66.7%. Trajectories shown in both panels results from the same set of simulations; simulations resulting in outbreak failure were discarded. Model parameters used for simulations in both panels: (*m, n*) = (6, 1), (1/*σ*, 1/*γ*) = (5.28 days, 2.9 days). Infection rate coefficient of hyper-spreaders was set *β*_h_ = 198 × *β*_n_ (where *β*_n_ is the infection rate coefficient for normal spreaders), which assures that in the deterministic limit 66.7% of infections are transmitted by hyper-spreaders. In turn *β*_n_ was set such that the average infection rate coefficient *β* = 2.97 × *β*_n_ gives 𝒯_d_ = 2 days (see equation (7) in Methods).

We estimated 𝒯_d_ in two ways: based on one month of growth of the number of new cases since the first registered case (‘30 days since the 1^st^ case’) and based on growth of new cases in the two-week period after the number of registered cases exceeds 100 (‘14 days since 100 cases’). As we are interested in the initial phase characterised by exponential growth, we assumed that the susceptible population remains constant. In figure 3 we show histograms of 𝒯_d_ calculated using either the ‘14 days since 100 cases’ method or the ‘30 days since the 1^st^ case’ method. One may observe that the histograms calculated using the ‘30 days since the 1^st^ case’ method are broader than those calculated using the ‘14 days since 100 cases’ method, and the width of all histograms increases with increasing infectiousness (which is set inversely proportional to *ρ*). When 𝒯_d_ is calculated using the ‘14 days since 100 cases’ method, its median value is slightly larger than 𝒯_d_ in the deterministic model (equal 2 days); however, when 𝒯_d_ is calculated using the ‘30 days since the 1^st^ case’ method, then for high infectiousness of super- and hyper-spreaders (correspondingly, for low *ρ*) its median value becomes much larger than the deterministic 𝒯_d_. Using the ‘30 days since the 1^st^ case’ method for the case of the lowest considered *ρ* = 1%, when super-spreaders (hyper-spreaders) have their infectiousness about 100 times (200 times) higher than the infectiousness of normal individuals, one obtains median 𝒯_d_ larger than 𝒯_d_ obtained in the deterministic model by 29% (67%), while for ‘14 days since 100 cases’ the 𝒯_d_ overestimation is negligible, 3% (6%). This difference is caused by low probability of appearance of super- or hyper-spreaders in the first weeks of the outbreak.

**Figure 3:**
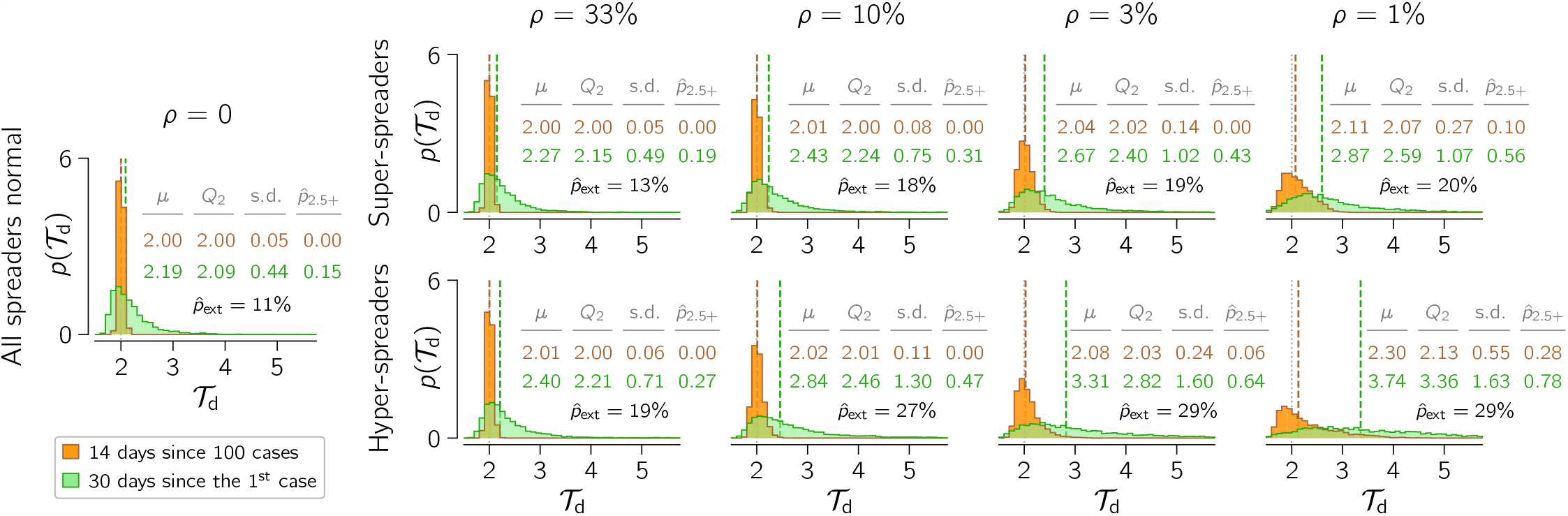
Estimation of the doubling time 𝒯_d_ based on stochastic simulations of the SEIR model with super- and hyper-spreaders. Histograms show probability density *p*(𝒯_d_) estimated using the ‘14 days since 100 cases’ method (orange) and the ‘30 days since the 1^st^ case’ method (green). In each column, *ρ* denotes a fixed proportion of super-spreaders (top row) or hyper-spreaders (bottom row) in the population. For decreasing proportions of super- and hyper-spreaders (from left, except the shared leftmost panel with *ρ* = 0, to right), their infection rate coefficient *β* has been reduced to give the same deterministic 𝒯_d_ = 2 days (vertical dotted grey lines). Remaining model parameters: (*m, n*) = (6, 1); (1/*σ*, 1/*γ*) = (5.28 days, 2.9 days). Each histogram results from 5,000 stochastic simulations starting from a single infected normal individual; trajectories resulting in outbreak failure were discarded; fraction of trajectories that resulted in epidemic extinction for given conditions is given as 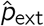. Each distribution is described in terms of its mean (*μ*), median (Q_2_ and vertical dashed lines), standard deviation (s.d.), and the fraction of probability mass for 𝒯_d_ > 2.5 days 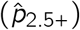.

Finally, we notice that 𝒯_d_ estimation for a given country based on available data is equivalent to the analysis of a single stochastic trajectory and that at a very initial stage the epidemic can cease. Probability of extinction is larger when a small fraction of super-spreaders is responsible for a large fraction of cases. In figure 3 we provided extinction probability, 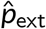, which in the extreme case of 1% of hyper-spreaders reaches 29%, whereas without hyper-spreaders (or super-spreaders) is 11%.

The examples shown in figure 2 and figure 3 are focused on the case in which the 𝒯_d_ = 2 days, which is close to 𝒯_d_ estimated for Spain and New York State. After removing super-spreaders (assumed to be responsible for 50% of transmissions) the doubling time would be equal 3.05 days, whereas after removing hyper-spreaders (responsible for 66.7% of transmissions) the doubling time would be equal to 4.24 days. The doubling times in the range 5.2–7.4, obtained by analysing early onsets of the epidemic (Wu *et al*. [14, 15] and Li *et al*. [16]), exceed our model prediction obtained after removing 66.7% of transmissions by hyper-spreaders, suggesting that the fraction of transmissions for which hyper-spreaders are responsible can be even larger. Endo *et al*. estimated that 80% of secondary transmissions could have been caused by 10% of infectious individuals [18].

## Conclusions

Based on epidemic data from China, United States, and six European countries, we have estimated that the basic reproduction number ℛ_0_ lies in the range 4.7–11.4 (5.6–7.3 for China), which is higher than most previous estimates [5, 8, 4]. There are two sources of the discrepancy in ℛ_0_ estimation. First, in agreement with data on the incubation period distribution (assumed to be the same as the latent period distribution), we used a model with six ‘exposed’ states, which substantially increases ℛ_0_(𝒯_d_) with respect to the models with one or two ‘exposed’ states. Second, we estimated 𝒯_d_ based on the two-week period of the exponential growth phase beginning on the day in which the number of cumulative registered cases exceeds 100, or when the number of cumulative registered fatalities exceeds 10. Importantly, values of 𝒯_d_ estimated from the growth of registered cases and from the growth of the registered fatalities led to similar ℛ_0_ estimates. This approach, in contrast to estimation of ℛ_0_ based on individual case reports, allows to implicitly take into account super-spreading events that substantially shorten 𝒯_d_. Spatial heterogeneity of the epidemic spread observed in many European countries, including Italy, Spain, and Germany, can be associated with larger or smaller super-spreading events that initiated outbreaks is particular regions of these countries.

Our estimates are consistent with current epidemic data in Italy, Spain, and France. As of April 24, 2020, these countries managed to terminate the exponential growth phase by means of country-wide quarantine. Current COVID-19 Community Mobility Reports [23] show about 80% reduction of mobility in retail and recreation, transit stations, and workplaces in these countries. Together with increased social distancing, this reduction possibly lowered the infection rate *β* at least five-fold; additionally, massive testing reduced the infectious period, 1/*γ*. Consequently, we suspect that the reproduction number *R* = *β*/*γ* was reduced more than five-fold, which brought it to the values somewhat smaller than 1. This suggests that ℛ_0_ in these countries could have been larger than 5.

## Methods

### SEIR model equations and parametrisation

The dynamics of our SEIR model is governed by the following system of ordinary differential equations:

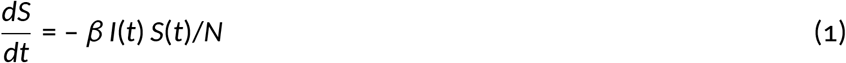

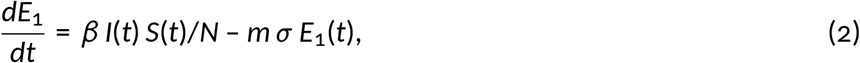

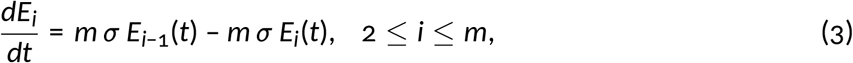

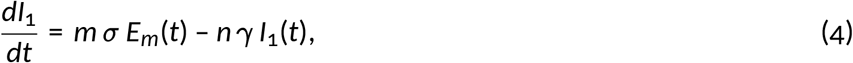

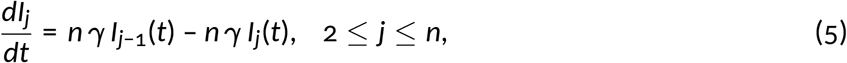

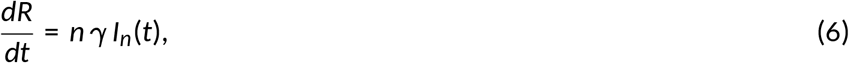

where *N* = *S*(*t*) + *E*_1_(*t*) + … + *E*_m_(*t*) + *I*_1_(*t*) + … + *I*_n_(*t*) + *R*(*t*) is the constant population size, and *I*(*t*) = *I*_1_(*t*) + … + *I*_n_(*t*) is the size of infectious subpopulation. As *m* is the number of ‘exposed’ substates and *n* is the number of ‘infectious’ substates, there are *m* + *n* + 2 equations in the system. In the early phase of the epidemic, 1 – *S*(*t*)/*N* ≪ 1 and with constant coefficients *β, σ*, and *γ* the growth of *R* (as well as *E*_*i*_ and *I*_*j*_) is exponential.

An important property of a given SEIR model parametrisation is its implied distribution of generation interval (GI), the period between subsequent infections events in a transmission chain. While the expected GI is easily computable from model parameters as 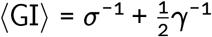 (the mean period of infectiousness is halved to reflect the assumption that the infection occurs in a random time during the period of infectiousness [17]), it can be hardly estimated based on even detailed epidemiological data. It should be noted that in some sources the formula ⟨G*I*⟩ = *σ*^−1^ + *γ*^−1^ is used (see, *e*.*g*., Ref. [24]), in which it is assumed that the infection occurs at the end of the period of infectiousness, not at a random point of this period. GI may be related to the serial interval, SI, the period between the occurrence of symptoms in the infector and the infectee. Although GI and SI may have different distributions, their means are expected to be equal and thus may be directly compared. Our parametrisation implies ⟨G*I*⟩ = 6.73 days, which is consistent with ⟨G*I*⟩ of the model by Ferguson *et al*. (6.5 days) [25] and the estimates of ⟨S*I*⟩ by Wu *et al*. (7.0 days) [15], Ma *et al*. (6.8 days) [26], or Bi *et al*. (6.3 days) [27].

The short period of effective infectiousness reflects the assumption that the individuals with confirmed infection are quickly isolated or self-isolated and then cannot infect other susceptible individuals. This enabled us to identify the reported increase of confirmed cases with the transfer of the individuals from the (last substate of the) ‘infectious’ compartment to the ‘removed’ compartment of the SEIR model. In addition to the currently diseased individuals that remain isolated, the ‘removed’ compartment contains the recovered (and assumed to be resistant) and deceased individuals.

We assume the same 1/*γ* = 2.9 days in all locations and times, being however aware that the mean infectious period may shorten over time due to the implementation of protective health care practices, increased diagnostic capacity, and contact tracing [27]. In turn, the mean latent period, 1/*σ*, may be considered an intrinsic property of the disease. As the distribution of the latent period is not known, as a simplification, in our model the distribution of the latent period (time since infection during which an infected individual cannot infect) is assumed to be the same as the distribution of the incubation period (time since infection during which an infected individual has not yet developed symptoms). We demonstrated the influence of 1/*σ* on the estimation of ℛ_0_ in electronic supplementary material (figure S1).

### Estimation of the doubling time and the basic reproduction number

Growth rates used for estimation of respective doubling times, 𝒯_d_, were determined by linear regression of the logarithm of the cumulative confirmed cases and cumulative deaths in the exponential phase of the epidemic separately in each of eight considered location. We discarded initial parts of trajectories with less than 100 confirmed cases (or 10 registered fatalities) and used two-week-long periods to strike a balance between: (*i*) analysis of epidemic progression when stochastic effects associated with individual transmission events, including super-spreading, are relatively small (see stochastic simulation trajectories in figure 2*a*) and (*ii*) analysis of the exponential phase of epidemic progression, which is relatively short due to imposition of restrictions. We expect that the trajectories of deaths may be less affected by under-reporting; nevertheless, doubling times obtained from growth rates of cumulative cases and cumulative deaths turn out to be quite consistent.

In the context of our SEIR model, the doubling time 𝒯_d_ and parameters *β, σ, γ, n, m* satisfy the relation

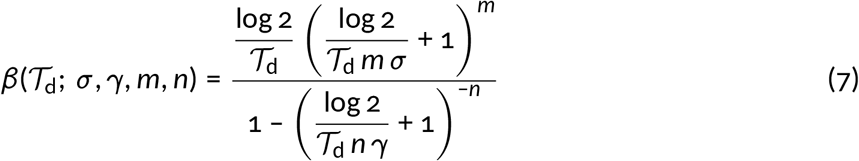

that enables calculation of the basic reproduction number using the doubling time 𝒯_d_ estimated directly from the epidemic data as

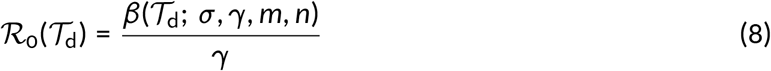

in accordance with Wearing *et al*. [11] and Wallinga & Lipsitch [12].

### Data accessibility

All data used in this theoretical study is referenced.

## Data Availability

All data are provided in manuscript and references therein

## Authors’ contributions

M.K. conceived study, performed model and data analysis, prepared figures and wrote manuscript; F.G. conceived study, performed model analysis and prepared figures; T.L. conceived study, wrote manuscript. All authors gave final approval for publication.

## ELECTRONIC SUPPLEMENTARY MATERIAL

**Figure S1:**
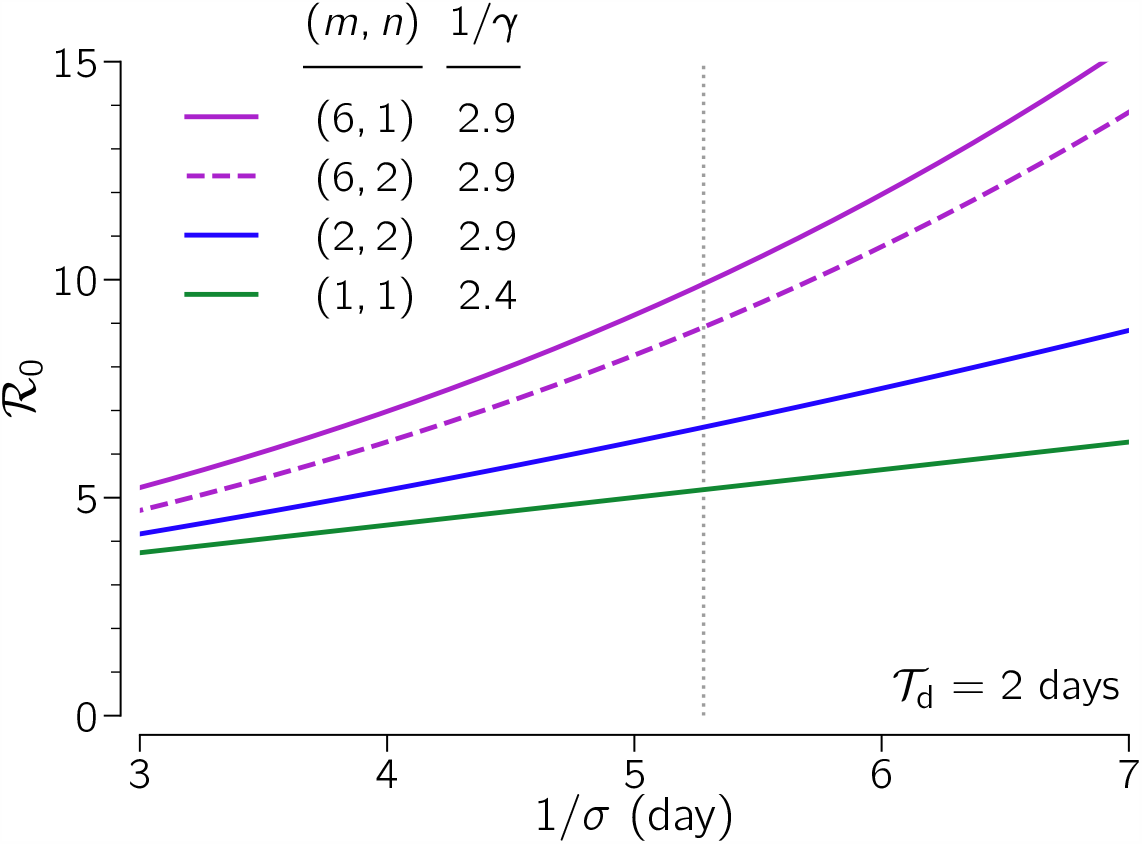
Basic reproduction number, ℛ_0_, *vs*. mean latent period, 1/*σ*. The SEIR model parameters are given in the legend; default 1/*σ* = 5.28 days is marked with a dotted vertical line. The assumed doubling time 𝒯_d_ is 2 days.

### Listing S1: BioNetGen language (BNGL)-encoded SEIR-type model of epi demic spread in the presence of super-spreaders

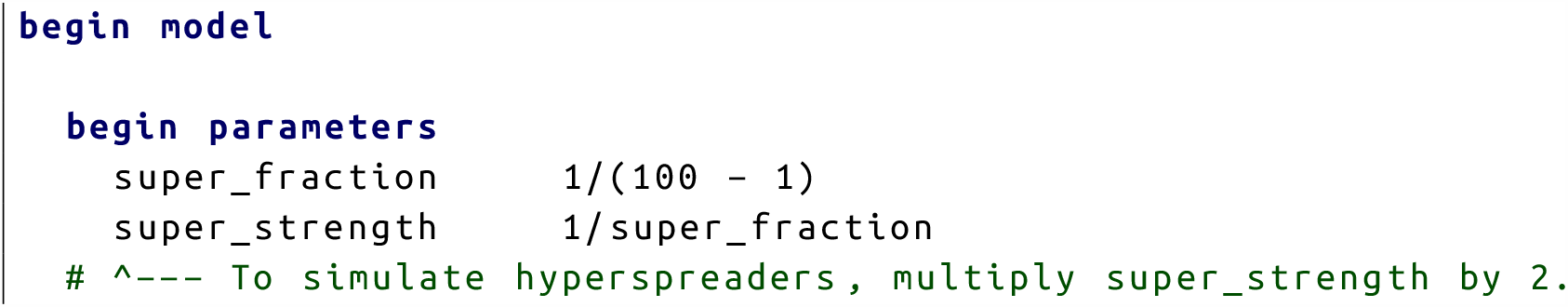
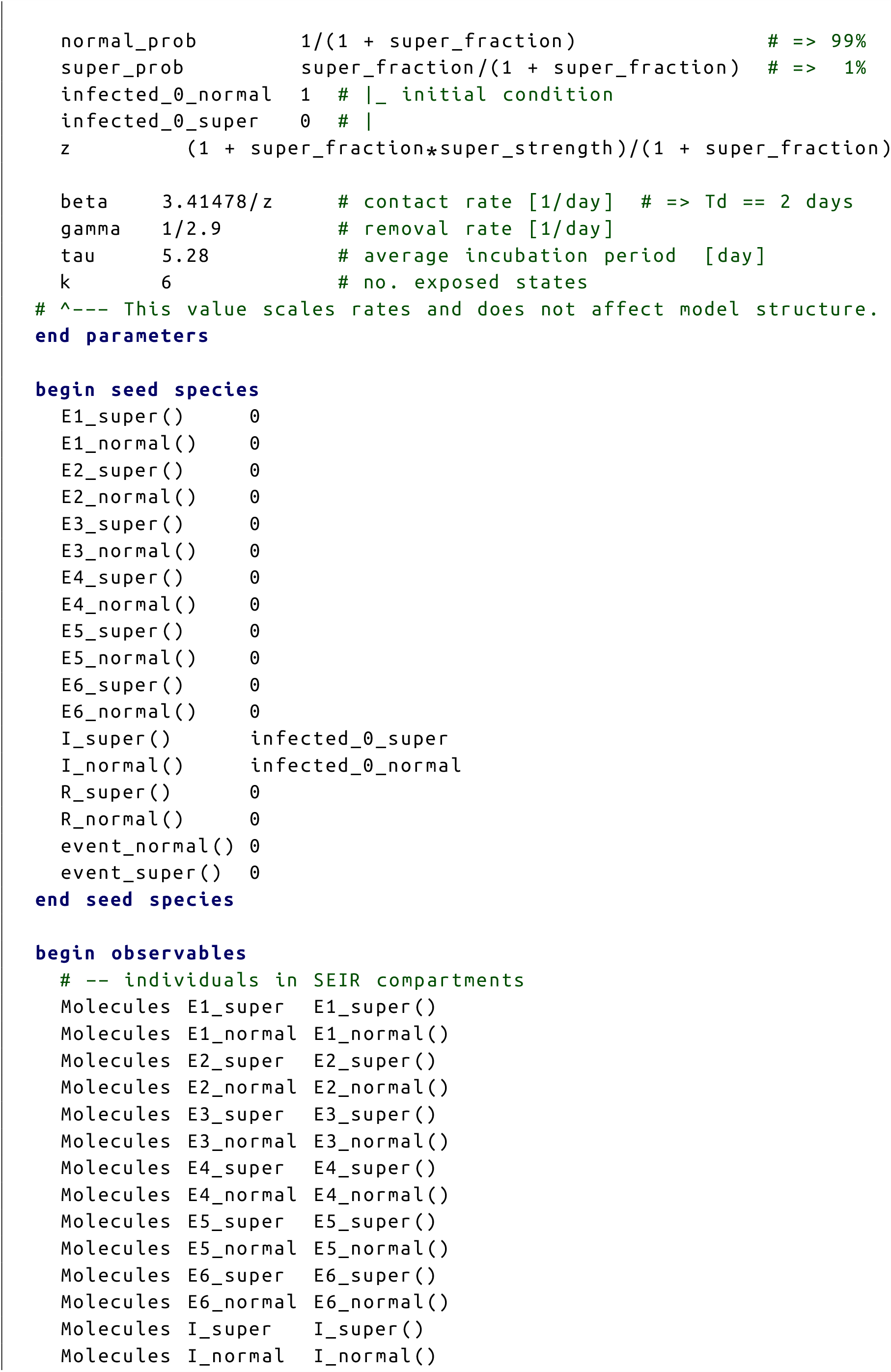
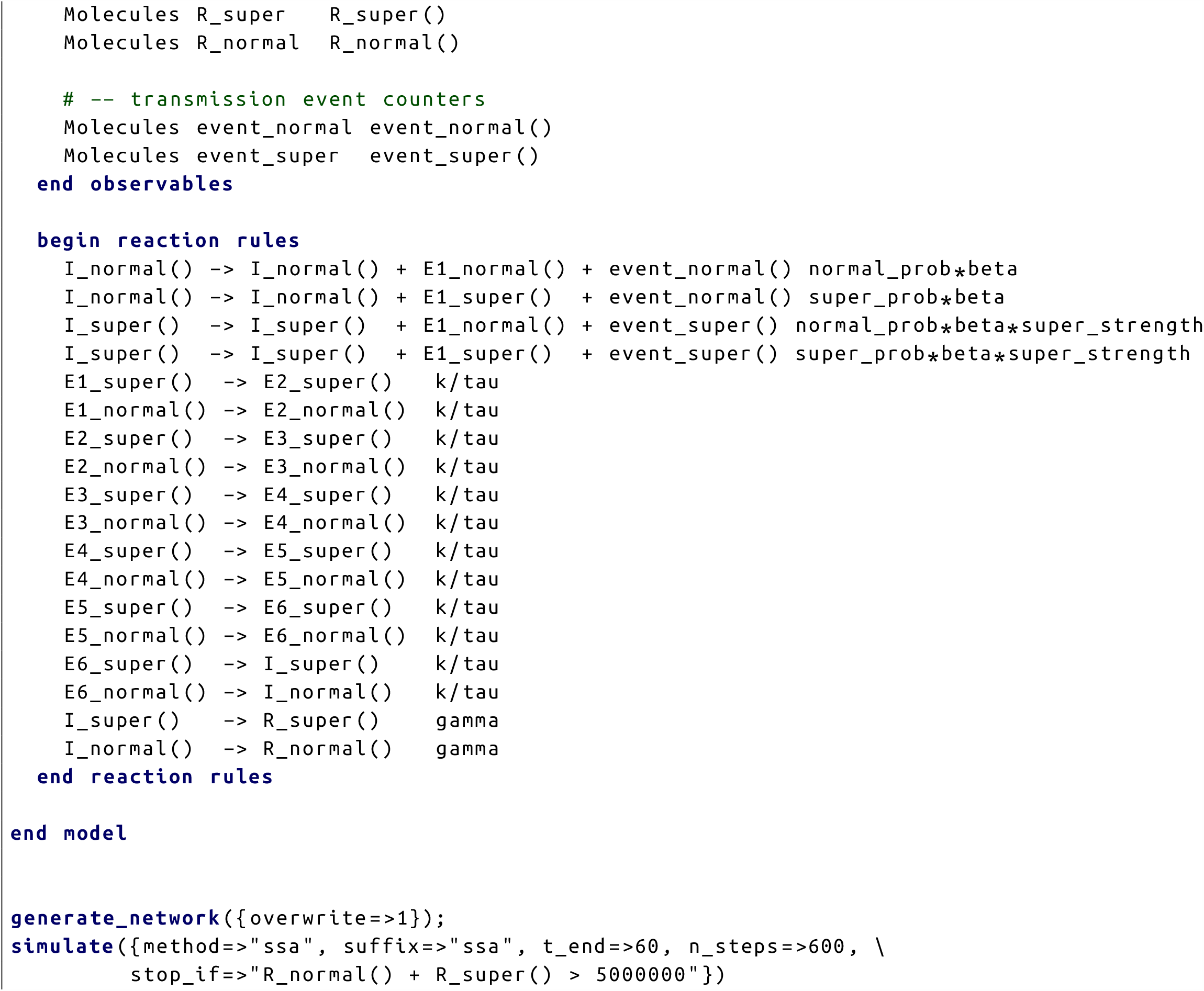

Parameters of the model are set so that the proportion of superspreaders in the population is set according to their infectiousness (relative w.r. to “normal” individuals) so that both groups, superspreaders and “normal” individuals, have identical total effective infectiousness. Also, contact rate is normalized so that the number of infecter “normal” individuals does not depend on the current relative infectiousness of superspreaders. The file contains both the model definition and, at the bottom, simulation protocol that runs a simulation with or without superspreaders. By changing `method⇒…`, one can run either a deterministic simulation (`method⇒ode`), using numerical integration of a resulting system of ODES, or an exact stochastic simulation (`method⇒ssa`) of a corresponding Markov chain, performed according the Gillespie algorithm. To simulate model dynamics, please download and install BioNetGen (from http://bionetgen.org). For the sake of convenience, you may use BioNetGen within RuleBender (http://www.rulebender.org). The model was developed in BioNetGen version 2.4.0 (in hope that it will be also compatible with future BioNetGen releases).

